# Behavioral determinants of preventive practices against German cockroach infestation among urban residents in Tehran, Iran

**DOI:** 10.64898/2026.07.04.26357085

**Authors:** Sajjad Moshavernia, Amrollah Azarm, Sara Bagherzade, Masoud Karimi, Haleh Ghaem Maralani, Mohammad Djaefar Moemenbellah-Fard

**Author notes:** **Corresponding Author**: Dr. M.D. **Moemenbellah-Fard** Research Center for Health Sciences, Institute of Health, Department of Biology and Control of Disease Vectors, School of Health, Shiraz University of Medical Sciences, Shiraz, Iran. **N.B.** Sajjad **Moshavernia** and Amrollah **Azarm** contributed equally to this work and should be considered co-first authors.

## Abstract

**Background:** German cockroach (*Blattella germanica*) infestation is an important urban environmental health menace associated with food contamination, allergic disease, and reduced quality of life. Long-term control depends not only on professional pest management, but also on residents’ knowledge and preventive behaviors. This study assessed the knowledge, Health belief model (HBM) constructs, self-efficacy, and preventive practices related to German cockroach infestation among urban residents in Tehran, Iran.

**Methods:** In this cross-sectional study, 120 adults with professionally confirmed household German cockroach infestation were recruited from licensed pest-control companies in Tehran. Data were collated using a 39-item HBM-based questionnaire assessing knowledge, perceived susceptibility, perceived severity, perceived benefits, perceived barriers, self-efficacy, and preventive practices. Descriptive statistics, Pearson correlation, and multiple linear regression were performed.

**Results:** Participants demonstrated modest knowledge regarding German cockroach biology (mean score: 0.538 ± 0.273) and moderate preventive practices (3.157 ± 1.006). Preventive practices were positively correlated with knowledge (r = 0.256, *P* = 0.005), perceived benefits (r = 0.292, *P* = 0.001), and self-efficacy (r = 0.244, *P* = 0.007). Regression analysis showed that the model explained 17.3% of the variance in preventive practices (R^2^ = 0.173, *P* = 0.001). Knowledge (β = 0.191, *P* = 0.036), perceived benefits (β = 0.231, *P* = 0.010), and self-efficacy (β = 0.229, *P* = 0.012) were significant predictors.

**Conclusions:** Urban residents with confirmed German cockroach infestation showed limited knowledge and moderate preventive behaviors. Knowledge, perceived benefits, and self-efficacy were independently associated with preventive practices and demonstrated modest predictive value. Interventions targeting these behavioral factors, alongside environmental and structural improvements, may enhance sustainable household cockroach control.

## Introduction

German cockroach infestation caused by *Blattella germanica* remains a major urban environmental health challenge, particularly in densely populated metropolitan areas [1]. Among domestic cockroach species, *B. germanica* is especially problematical due to its high reproductive capacity, strong adaptation to indoor environments, cryptic harboring behavior, and rapid re-infestation potential [2]. Beyond being a nuisance, infestations are associated with food contamination, mechanical transmission of pathogens, allergic sensitization, asthma exacerbation, psychological stress, and reduced quality of life [3].

Despite widespread insecticide use, effective long-term management of German cockroach infestation remains difficult due to insecticide resistance, hidden harborages, poor sanitation, structural deterioration, and re-infestation from adjacent apartments [4]. These challenges have shifted pest-management strategies toward integrated pest management (IPM), which combines sanitation, environmental modification, exclusion, monitoring, and targeted pesticide application [5], particularly under the One Health initiative approach. However, the effectiveness of IPM depends heavily on residents’ behaviors, as household practices directly influence food availability, moisture, clutter, and harboring opportunities for cockroaches [6].

Behavioral determinants are therefore critical for long-term infestation control [7]. Residents’ knowledge of cockroach biology and infestation pathways may influence recognition of infestation sources and adoption of preventive practices [8]. Misconceptions regarding harboring sites or transmission routes may lead to ineffective control behaviors and recurrent infestation. In addition to knowledge, psychological factors may also influence preventive actions [9].

The Health Belief Model (HBM) is a widely used framework for explaining preventive health behaviors and includes perceived susceptibility, perceived severity, perceived benefits, perceived barriers, and self-efficacy [10]. Although HBM has been extensively applied in environmental and vector-control research, its application to household cockroach management remains limited [11].

Although household cockroach infestation has been investigated in multiple environmental health studies, prior research has primarily focused on infestation prevalence, insecticide resistance, or general sanitation practices rather than behavioral determinants of household control [4]. Moreover, many previous studies relied on self-reported infestation status, which increases the risk of exposure misclassification [6]. To our knowledge, few studies have applied the Health Belief Model to evaluate behavioral determinants of preventive practices specifically among residents with professionally confirmed German cockroach infestation. Addressing this gap may provide more accurate insights into the cognitive and behavioral factors influencing sustainable household infestation control [12].

This study, therefore, aimed to assess the knowledge, HBM constructs, self-efficacy, and preventive practices related to German cockroach infestation among urban residents in the Iranian capital city, Tehran, with professionally confirmed household infestation. We hypothesized that greater knowledge, stronger perceived benefits, lower perceived barriers, and higher self-efficacy would be associated with stronger preventive measures.

## Materials and Methods

### Study Design and Settings

This cross-sectional analytical study was conducted between January and June 2026 in Tehran, Iran, to investigate knowledge, Health Belief Model (HBM) constructs, self-efficacy, and preventive practices related to household infestation by the German cockroach (*Blattella germanica*). The study was designed and reported in accordance with the Strengthening the Reporting of Observational Studies in Epidemiology (STROBE) recommendations for cross-sectional studies [13].

### Study Population and Eligibility Criteria

The study population consisted of adults aged 18 years or older residing in Tehran who sought professional pest-control services due to suspected household cockroach infestation during the study period. Participants were eligible if they were at least 18 years old, lived in Tehran, had active household infestation with *Blattella germanica* confirmed by professional inspection, and provided written informed consent. Individuals were excluded if they submitted incomplete questionnaires with substantial missing responses, declined participation or withdrew consent, or had previously participated in the study.

### Infestation Confirmation and Species Identification

Infestation confirmation was conducted by two certified pest-management inspectors with formal training in medical entomology using a standardized inspection checklist. Each household underwent a structured 15–20-minute visual inspection covering kitchens, bathrooms, sewage access points, wall cracks, furniture crevices, and food storage areas. Inspectors recorded evidence of active infestation including live cockroaches, ootheca, fecal spotting, and harboring signs. Species identification of *Blattella germanica* was confirmed using standard morphological characteristics, including body size, light-brown coloration, and two longitudinal dark stripes on the pronotum [14]. Disagreements in identification were resolved through consensus review.

### Sampling Strategy and Sample Size

Participants were recruited using consecutive sampling from clients of a licensed pest-control company serving northern, southern, eastern, and western districts of Tehran. Consecutive recruitment was adopted to minimize investigator-driven selection bias and improve representativeness across diverse urban settings. Sample size was estimated a priori using G*Power version 3.1 for multiple linear regression analysis with six predictors [15]. Assuming a medium effect size (f^2^ = 0.15), 80% statistical power, and a two-sided alpha level of 0.05, the minimum required sample size was estimated at 92 participants. To compensate for potential non-response and incomplete questionnaires, oversampling was planned. Of 146 eligible individuals approached, 120 completed the questionnaire and were included in the final analysis, yielding a response rate of 82.2%.

### Questionnaire Development and Measurement

Data were collated using a structured questionnaire specifically developed for this study based on literature review, expert consultation, and the theoretical framework of the Health Belief Model [16]. Questionnaire development included literature synthesis on German cockroach biology, integrated pest management, and behavioral determinants of household pest control, followed by item generation, expert review, and pilot refinement for clarity and comprehension. The final instrument contained 39 items across seven domains: knowledge (16 items), perceived susceptibility (3 items), perceived severity (4 items), perceived benefits (4 items), perceived barriers (3 items), self-efficacy (4 items), and preventive practices (5 items).

Knowledge items were designed to assess participants’ understanding of German cockroach biology, infestation pathways, harboring sites, and common misconceptions frequently encountered during pest-control consultations. Knowledge items were scored dichotomously, with correct responses assigned a score of 1 and incorrect or “do not know” responses assigned 0. A composite knowledge score was calculated by averaging item scores, resulting in values ranging from 0 to 1, with higher values indicating greater knowledge.

HBM constructs and self-efficacy were assessed using five-point Likert scales. Perceived susceptibility, perceived severity, and perceived barriers were measured on scales ranging from 1 (strongly disagree) to 5 (strongly agree). Perceived benefits and self-efficacy were measured on scales ranging from 1 (very low) to 5 (very high). Preventive practices were measured using a five-point frequency scale ranging from 1 (never/very low) to 5 (always/very high). Composite scores for each domain were calculated as arithmetic means of their respective items, with higher scores reflecting stronger endorsement of the construct or more frequent engagement in preventive behavior.

### Validity and Reliability Assessment

Content validity was assessed by an expert panel consisting of ten specialists, including four medical entomologists, three environmental health specialists, two health education experts, and one epidemiologist. Each expert independently evaluated questionnaire items for relevance, clarity, simplicity, and necessity. Content Validity Ratio (CVR) was calculated according to Lawshe’s method, and Content Validity Index (CVI) was calculated using item relevance ratings. Items failing to meet recommended thresholds were revised or removed. The final questionnaire demonstrated excellent content validity, with an overall CVR of 0.89 and CVI of 0.92 [17].

Although content validity and internal consistency were satisfactory, construct validity was not evaluated using exploratory or confirmatory factor analysis [18]. Therefore, the latent structure of the questionnaire domains should be interpreted cautiously and requires further psychometric validation in future studies.

### Data Collection Procedure

Eligible participants were invited to participate after confirmation of household infestation. Following written informed consent, participants completed the questionnaire either independently or with assistance from trained research personnel when clarification was needed. Standardized instructions were provided to all participants to minimize information bias and ensure consistency in data collection. All responses were anonymized prior to statistical analysis.

### Statistical Analysis

Data were analyzed using IBM SPSS Statistics version 26. Descriptive statistics were used to summarize demographic characteristics and questionnaire responses, including means, standard deviations, medians, frequencies, and percentages as appropriate. Distributional properties of continuous variables were assessed using histograms, Q–Q plots, skewness and kurtosis indices, and Shapiro–Wilk testing when necessary.

Associations between demographic characteristics and study outcomes were examined using independent-samples t-tests for binary variables, one-way analysis of variance (ANOVA) for categorical variables with more than two groups, and Pearson correlation analysis for continuous variables. Pearson correlation coefficients were further calculated to evaluate relationships among knowledge, HBM constructs, self-efficacy, and preventive practices.

Multiple linear regression analysis was performed to identify independent predictors of preventive practices [19]. Knowledge, perceived susceptibility, perceived severity, perceived benefits, perceived barriers, and self-efficacy were entered as independent variables, while preventive practice score served as the dependent variable. Regression assumptions including linearity, independence of residuals, normality of residuals, homoscedasticity, and multicollinearity were assessed prior to model interpretation [20]. Multicollinearity was evaluated using variance inflation factor (VIF) and tolerance statistics. VIF values below 5 and tolerance values above 0.20 were considered acceptable. In the final model, all VIF values were below 2.0, indicating no evidence of problematic multicollinearity. All statistical tests were two-sided, and *P*-values less than 0.05 were considered significant.

Regression diagnostics included visual inspection of residual plots, assessment of standardized residual normality using Q–Q plots, evaluation of independence using the Durbin–Watson statistic, and examination of homoscedasticity using residual-versus-fitted plots. No substantial violations of regression assumptions were detected.

### Bias Control and Ethical Considerations

Several measures were implemented to minimize potential bias. Objective confirmation of infestation reduced exposure misclassification, standardized data collection procedures minimized interviewer-related bias, and consecutive sampling reduced selective recruitment [21]. Nevertheless, since participants were recruited from pest-control service users, some selection bias toward individuals with greater infestation severity or higher health awareness could not be excluded.

The study protocol was approved by the Ethics Committee of Shiraz University of Medical Sciences (Approved Ethics Code: IR.SUMS.SCHEANUT.REC.1405.013). The study was conducted in accordance with the Declaration of Helsinki [22]. Written informed consent was obtained from all participants before enrollment, and all collated data were anonymized and handled confidentially throughout the study.

## Results

### Participant Characteristics

Among 146 eligible individuals approached during the study period, 120 completed the questionnaire and were included in the final analyses, yielding a response rate of 82.2% (Figure 1). No missing values were observed in demographic variables or questionnaire responses; therefore, complete-case analysis was performed. The mean age of participants was 30.46 ± 5.09 years (median: 30.0 years). Of all participants, 61 (50.8%) were female and 59 (49.2%) were male. Most participants held a bachelor’s degree (n = 71, 59.2%), followed by diploma-level education (n = 34, 28.3%). Regarding income level, 69 participants (57.5%) reported middle income, 36 (30.0%) upper-middle income, 13 (10.8%) high income, and 2 (1.7%) low income. The participant recruitment and eligibility process is represented in Figure 1, and demographic characteristics are summarized in **Table** 1.

**Figure 1.**
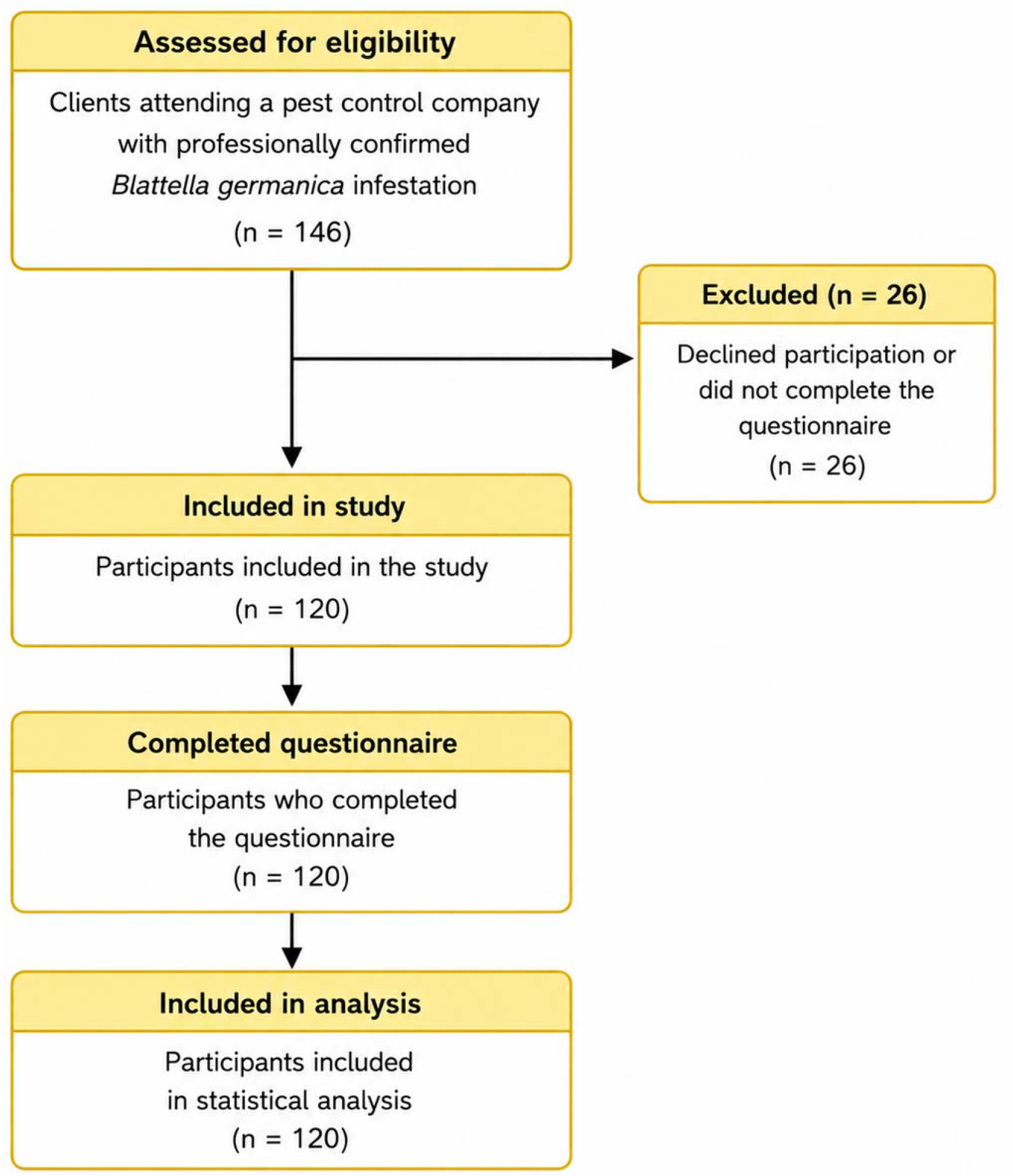
STROBE-compliant participant flow diagram showing recruitment, eligibility assessment, exclusions, and final analytic sample.

**Table 1.** Demographic characteristics of study participants with professionally confirmed household German cockroach infestation in Tehran, Iran (n = 120).

| Characteristic | Category | n (%) / Mean ± SD |
| --- | --- | --- |
| Age | Years | 30.46 ± 5.09 |
| Sex | Female | 61 (50.8) |
|  | Male | 59 (49.2) |
| **Education** | High school | 3 (2.5) |
|  | Diploma | 34 (28.3) |
|  | Bachelor | 71 (59.2) |
|  | Master | 9 (7.5) |
|  | PhD | 3 (2.5) |
| **Income** | Low | 2 (1.7) |
|  | Middle | 69 (57.5) |
|  | Upper-middle | 36 (30.0) |
|  | High | 13 (10.8) |

### Descriptive Statistics of Study Variables

Participants demonstrated suboptimal knowledge regarding German cockroach biology and infestation pathways, with a mean knowledge score of 0.538 ± 0.273. Mean scores for perceived susceptibility, perceived severity, perceived benefits, perceived barriers, self-efficacy, and preventive practices were 3.044 ± 1.072, 3.229 ± 0.931, 3.429 ± 0.962, 2.931 ± 1.011, 3.081 ± 1.065, and 3.157 ± 1.006, respectively, indicating mid-range across most Health Belief Model constructs and preventive behaviors. Internal consistency reliability was acceptable to excellent across all questionnaire domains, with Cronbach’s alpha coefficients ranging from 0.764 to 0.860. Detailed descriptive statistics and reliability estimates for all study variables are presented in **Table 2**. The highest correct response rates were observed for items related to flight ability (75.0%) and jumping ability (72.5%), whereas the lowest correct response rates were observed for misconceptions regarding reproduction on the human body (27.5%) and reproduction in clothes or fabrics (30.8%). Internal consistency reliability was assessed using Cronbach’s alpha coefficients for each questionnaire domain. Cronbach’s alpha values ≥0.70 were considered acceptable.

**Table 2.** Descriptive statistics and internal consistency reliability of knowledge, Health Belief Model constructs, self-efficacy, and preventive practice domains.

| **Construct** | **Items** | **Cronbach $\alpha$** | **Mean ± SD** | **Range** |
| --- | --- | --- | --- | --- |
| **Knowledge** | 16 | 0.860 | $0.538 \pm 0.273$ | 0–1 |
| **Susceptibility** | 3 | 0.822 | $3.044 \pm 1.072$ | 1–5 |
| **Severity** | 4 | 0.819 | $3.229 \pm 0.931$ | 1–5 |
| **Benefits** | 4 | 0.825 | $3.429 \pm 0.962$ | 1–5 |
| **Barriers** | 3 | 0.764 | $2.931 \pm 1.011$ | 1–5 |
| **Self-efficacy** | 4 | 0.854 | $3.081 \pm 1.065$ | 1–5 |
| **Practices** | 5 | 0.859 | $3.157 \pm 1.006$ | 1–5 |

Cronbach's $\alpha$ values $\geq 0.70$ indicated acceptable internal consistency.

### Demographic Associations with Knowledge and Preventive Practices

No significant sex differences were observed in knowledge scores (t = 0.565, p = 0.573) or preventive practice scores (t = 1.181, p = 0.240). Educational level showed a significant association with both knowledge and preventive practices. Participants with higher educational attainment demonstrated significantly higher knowledge scores (F = 25.649, p < 0.001) and stronger preventive practices (F = 3.314, p = 0.013). Age was not significantly associated with knowledge (r = 0.102, p = 0.267) or preventive practices (r = 0.091, p = 0.323). Detailed results are presented in **Table 3**. Participants with higher educational attainment generally demonstrated greater knowledge regarding German cockroach biology and reported stronger preventive practices compared with those having lower educational levels.

**Table 3.** Associations between demographic characteristics and knowledge and preventive practice scores.

| Variable | Outcome | Test | Statistic | Effect Size | p-value |
| --- | --- | --- | --- | --- | --- |
| Sex | Knowledge | Independent t-test | $t = 0.565$ | Cohen's $d = 0.10$ | 0.573 |
| Sex | Preventive Practices | Independent t-test | $t = 1.181$ | Cohen's $d = 0.22$ | 0.240 |
| Education | Knowledge | One-way ANOVA | $F = 25.649$ | $\eta^2 = 0.47$ | <0.001 |
| Education | Preventive Practices | One-way ANOVA | $F = 3.314$ | $\eta^2 = 0.10$ | 0.013 |
| Age | Knowledge | Pearson correlation | $r = 0.102$ | — | 0.267 |
| Age | Preventive Practices | Pearson correlation | $r = 0.091$ | — | 0.323 |

**Notes:** Cohen's $d$ values of 0.2, 0.5, and 0.8 indicate small, moderate, and large effect sizes, respectively. $\eta^2$ values of 0.01, 0.06, and 0.14 indicate small, medium, and large effects.

### Knowledge Regarding German Cockroach Biology and Infestation Pathways

Analysis of individual knowledge items revealed several misconceptions regarding German cockroach biology and infestation routes. Correct response rates ranged from 27.5% to 75.0%. The highest proportion of correct responses was observed for the item regarding flight ability (75.0%), followed by jumping ability (72.5%) and reproduction in bathrooms/toilets (70.0%). In contrast, substantial misconceptions were observed regarding reproduction on the human body (27.5%), reproduction in clothes and fabrics (30.8%), and reproduction in bedding materials (39.2%). Awareness of dispersal pathways through second-hand furniture, sewage systems, and neighboring apartments was moderate, with correct response rates ranging from 57.5% to 65.0%. Detailed item-level results are shown in **Table 4**.

**Table 4.**
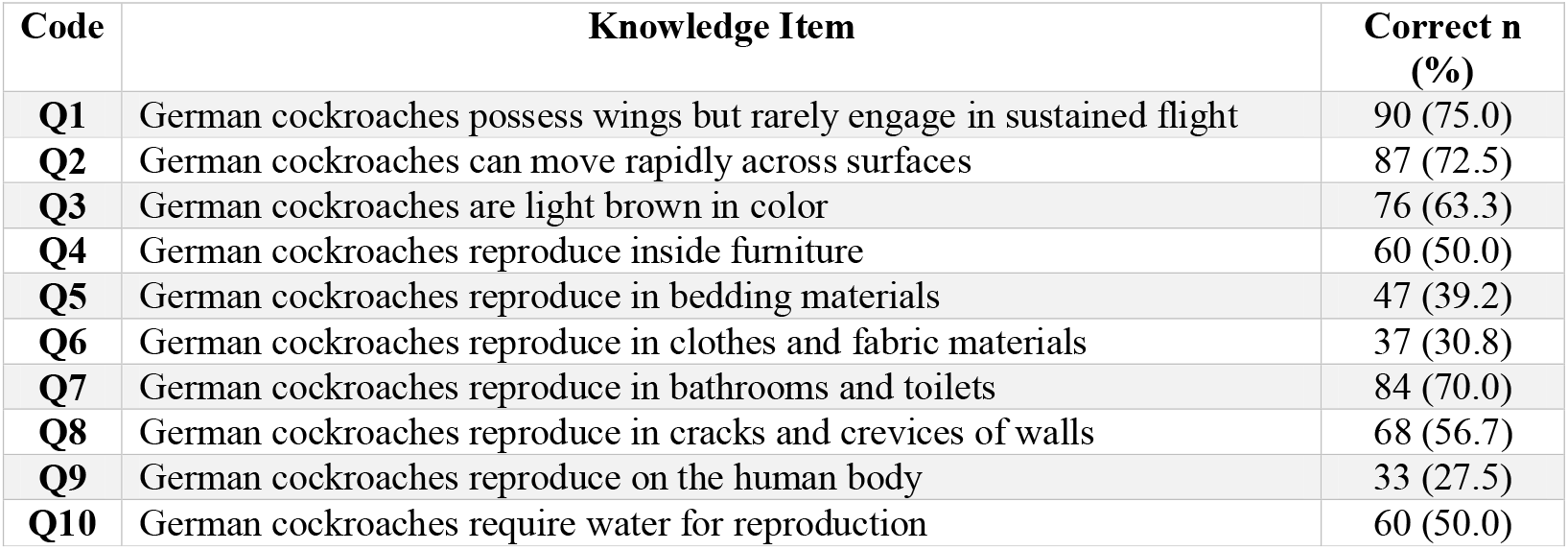
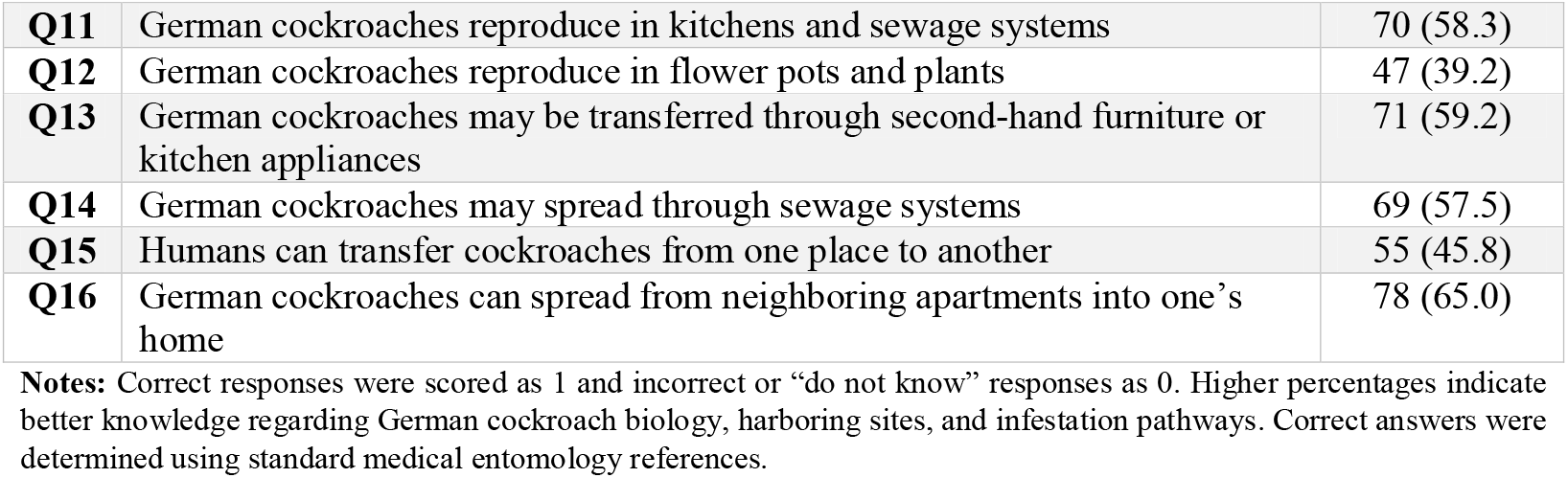
Correct response rates for knowledge items regarding German cockroach biology and infestation pathways among urban residents in Tehran, Iran (n = 120).

### Health Belief Model Constructs

Participants reported moderate perceived susceptibility to German cockroach infestation. The highest susceptibility score was related to building age as a contributing factor (mean = 3.275 ± 1.223), whereas sanitation-related susceptibility showed slightly lower perceived risk (mean = 2.792 ± 1.321). Perceived severity was also moderate. Participants most strongly agreed that cockroaches could contaminate food (mean = 3.683 ± 1.130) and transmit diseases (mean = 3.508 ± 1.077), while perceived social embarrassment due to infestation was comparatively lower (mean = 3.125 ± 1.254).

Perceived benefits of preventive measures were generally moderate to high. Household cleaning and sanitation were perceived as the most beneficial control measures (mean = 3.725 ± 1.084), followed by insecticide spraying (mean = 3.525 ± 1.195) and poisoned bait use (mean = 3.367 ± 1.276). Discarding old household items received comparatively lower benefit scores (mean = 3.100 ± 1.191).

Perceived barriers were moderate overall. Time burden associated with cockroach control showed the highest barrier score (mean = 3.033 ± 1.236), followed by cost of control products (mean = 2.983 ± 1.202) and professional pest-control expenses (mean = 2.775 ± 1.240). Self-efficacy scores were moderate, with the highest confidence reported for willingness to discard unnecessary household items (mean = 3.308 ± 1.275), whereas confidence in affording professional pest-control services was lowest (mean = 2.783 ± 1.271). Detailed item-level responses for perceived susceptibility, severity, benefits, barriers, and self-efficacy are presented in Supplementary Tables S1–S5.

### Preventive and Control Practices

Preventive practice scores indicated moderate adoption of household infestation-control behaviors. Regular household cleaning was the most frequently reported preventive behavior (mean = 3.475 ± 1.243), followed by insecticide spraying (mean = 3.375 ± 1.174) and poisoned bait use (mean = 3.150 ± 1.281). Professional consultation with pest-control specialists was less frequent (mean = 2.975 ± 1.233), and discarding old household items was the least frequently reported preventive behavior (mean = 2.808 ± 1.349). Item-level preventive practice responses are summarized in Supplementary Table S6.

### Correlation Analysis

Pearson correlation analysis identified significant positive correlations between preventive practices and knowledge (r = 0.256, p = 0.005), perceived benefits (r = 0.292, p = 0.001), and self-efficacy (r = 0.244, p = 0.007). No significant correlations were observed between preventive practices and perceived susceptibility (r = −0.031, p > 0.05), perceived severity (r = −0.024, p > 0.05), or perceived barriers (r = −0.012, p > 0.05). Among predictor variables, knowledge showed a positive correlation with perceived benefits (r = 0.237, p < 0.01), whereas perceived barriers demonstrated a negative correlation with self-efficacy (r = −0.206, p < 0.05). The full correlation matrix is presented in **Table 5**. Although significant, the observed correlations were small-modest in magnitude, suggesting that preventive practices are influenced by additional factors beyond the measured behavioral constructs.

**Table 5.** Pearson correlation matrix among knowledge, Health Belief Model constructs, self-efficacy, and preventive practices.

| Variable | 1 | 2 | 3 | 4 | 5 | 6 | 7 |
| --- | --- | --- | --- | --- | --- | --- | --- |
| **1 Knowledge** | 1 |  |  |  |  |  |  |
| **2 Susceptibility** | 0.121 | 1 |  |  |  |  |  |
| **3 Severity** | -0.141 | 0.129 | 1 |  |  |  |  |
| **4 Benefits** | 0.237** | 0.000 | -0.022 | 1 |  |  |  |
| **5 Barriers** | 0.021 | -0.098 | -0.012 | -0.076 | 1 |  |  |
| **6 Self-efficacy** | 0.116 | 0.155 | -0.110 | 0.086 | -0.206* | 1 |  |
| **7 Practices** | 0.256** | -0.031 | -0.024 | 0.292** | -0.012 | 0.244** | 1 |

Notes: Pearson correlation coefficients are shown. \* $p < 0.05$ ; \*\* $p < 0.01$ (two-tailed).

### Multiple Linear Regression Analysis

Pearson correlation analysis identified significant positive correlations between preventive practices and knowledge (r = 0.256, p = 0.005), perceived benefits (r = 0.292, p = 0.001), and self-efficacy (r = 0.244, p = 0.007), whereas no significant correlations were observed with perceived susceptibility, perceived severity, or perceived barriers. The full correlation matrix among knowledge, Health Belief Model constructs, self-efficacy, and preventive practices is presented in **Table 5**. Among significant predictors, perceived benefits showed the strongest standardized association with preventive practices (β = 0.231), followed closely by self-efficacy (β = 0.229) and knowledge (β = 0.191). Although significant, the model demonstrated modest explanatory power, indicating that a substantial proportion of variance in preventive practices remained unexplained by the included predictors.

Multiple linear regression analysis was subsequently performed to identify independent predictors of preventive practices, and the regression coefficients are represented in **Table 6**. The overall regression model was significant (R = 0.416, R^2^ = 0.173, adjusted R^2^ = 0.129, F (6,113) = 3.948, p = 0.001), indicating that the included predictors collectively explained 17.3% of the variance in preventive practice scores. Knowledge was a significant positive predictor of preventive practices (B = 0.704, SE = 0.333, β = 0.191, t = 2.117, p = 0.036, 95% CI: 0.045–1.364). Perceived benefits also showed a significant positive association with preventive practices (B = 0.241, SE = 0.093, β = 0.231, t = 2.605, p = 0.010, 95% CI: 0.058–0.425). Similarly, self-efficacy was independently associated with stronger preventive practices (B = 0.217, SE = 0.085, β = 0.229, t = 2.562, p = 0.012, 95% CI: 0.049–0.384). In contrast, perceived susceptibility (β = −0.092, p = 0.301), perceived severity (β = 0.046, p = 0.606), and perceived barriers (β = 0.041, p = 0.646) were not significant predictors in the multivariable model.

**Table 6.** Multiple linear regression analysis predicting preventive practices among participants with household German cockroach infestation.

| Predictor | B | SE | $\beta$ | t | p-value | 95% CI for B | VIF |
| --- | --- | --- | --- | --- | --- | --- | --- |
| **Knowledge** | 0.704 | 0.333 | 0.191 | 2.117 | 0.036 | 0.045 to 1.364 | 1.113 |
| **Susceptibility** | -0.086 | 0.083 | -0.092 | -1.038 | 0.301 | -0.251 to 0.078 | 1.072 |
| **Severity** | 0.049 | 0.095 | 0.046 | 0.518 | 0.606 | -0.139 to 0.238 | 1.059 |
| **Benefits** | 0.241 | 0.093 | 0.231 | 2.605 | 0.010 | 0.058 to 0.425 | 1.072 |
| **Barriers** | 0.040 | 0.088 | 0.041 | 0.460 | 0.646 | -0.133 to 0.214 | 1.058 |
| **Self-efficacy** | 0.217 | 0.085 | 0.229 | 2.562 | 0.012 | 0.049 to 0.384 | 1.096 |

**Model statistics:** $R = 0.416$ , $R^2 = 0.173$ , Adjusted $R^2 = 0.129$ , $F(6,113) = 3.948$ , $p = 0.001$ .
**Notes:** B = unstandardized regression coefficient; SE = standard error; $\beta$ = standardized regression coefficient; CI = confidence interval; VIF = variance inflation factor.

Regression diagnostics indicated no substantial violations of model assumptions. Variance inflation factor values ranged from 1.058 to 1.113 and tolerance values ranged from 0.899 to 0.945, confirming the absence of problematic multicollinearity.

## Discussion

This study examined knowledge, Health Belief Model (HBM) constructs, self-efficacy, and preventive practices related to household infestation by *Blattella germanica* among urban residents in Tehran with professionally confirmed infestation. Three main findings emerged. First, participants demonstrated suboptimal knowledge regarding infestation pathways. Second, knowledge, perceived benefits, and self-efficacy were positively associated with preventive practices. Third, perceived susceptibility, perceived severity, and perceived barriers were not significant predictors after multivariable adjustment.

Participants showed substantial misconceptions regarding German cockroach ecology, particularly concerning harboring sites and infestation pathways [23]. Limited knowledge may contribute to ineffective control behaviors, including overreliance on insecticide spraying while neglecting essential environmental measures such as sanitation, moisture reduction, and crack sealing [24]. Since integrated pest management (IPM) depends heavily on environmental control, inadequate knowledge may directly undermine sustainable infestation management [25]. Correcting common misconceptions may be associated with improved household control behaviors, although causal relationships cannot be established from cross-sectional data [26]. However, due to the cross-sectional design, causal relationships cannot be inferred.

Perceived benefits emerged as one of the strongest predictors of preventive practices [27]. Participants who believed sanitation, bait use, and environmental management were effective reported stronger preventive behaviors. This finding supports the HBM assumption that perceived utility of interventions strongly influences behavioral adoption [28]. Educational interventions should therefore emphasize not only factual knowledge but also the practical effectiveness of recommended control measures [29].

Self-efficacy was also a significant predictor, highlighting the importance of confidence in performing control behaviors. Cockroach management requires sustained effort and behavioral consistency [30]. Individuals who feel capable of implementing preventive measures may be more likely to maintain long-term control practices [31]. Practical skill-based interventions may support adherence to preventive behaviors, although intervention studies are needed to confirm effectiveness [32].

In contrast, perceived susceptibility, severity, and barriers were not significantly associated with preventive practices. This suggests that risk awareness alone may be insufficient to drive behavior change in persistent urban infestations [33]. Residents may perceive cockroach infestation as unavoidable because of structural factors such as aging buildings, shared plumbing systems, and re-infestation from neighboring apartments [34]. Under such conditions, even high perceived risk may not translate into action [35].

Educational attainment was positively associated with both knowledge and preventive practices, indicating that educational inequalities may contribute to disparities in household pest management [36]. Targeted educational strategies may therefore be especially beneficial for populations with lower educational attainment [37].

A major strength of this study was the objective confirmation of infestation through professional household inspection, reducing exposure misclassification common in self-reported studies [38]. In addition, the study applied a theory-based behavioral framework with acceptable psychometric properties [39].

Several limitations should be considered. The cross-sectional design precludes causal inference [40]. Recruitment from clients of a licensed pest-control company may have introduced substantial selection bias. Individuals seeking professional pest-control services may differ systematically from the general population in terms of socioeconomic status, infestation severity, health awareness, and willingness to invest in household control measures [41]. Consequently, the study sample may overrepresent households with greater resources or more severe infestations, thereby limiting external validity and generalizability to the broader urban population of Tehran. Preventive practices were assessed using self-reported responses rather than objective household observations [42]. This may have introduced recall bias and social desirability bias, as participants may have overreported desirable behaviors such as cleaning, bait use, or sanitation practices [43]. Several potentially important confounding variables were not included in the regression model, including household income, housing type, building age, household size, sanitation quality, and infestation severity. These factors may influence both behavioral perceptions and preventive practices and could partially explain the modest explanatory power of the model [44].

The regression model explained only 17.3% of variance in preventive practices, indicating that HBM constructs alone incompletely explain behavior. Household cockroach control is likely influenced by broader structural, environmental, and socioeconomic determinants [45]. Future studies should integrate behavioral and ecological variables to better understand infestation-control behavior.

Overall, interventions focused solely on risk communication may have limited effectiveness. Sustainable cockroach control likely requires combined behavioral education, environmental sanitation, housing improvements, and accessible professional pest-management services.

## Conclusions

This study provides novel evidence regarding the behavioral determinants of household German cockroach (*Blattella germanica*) control among urban residents in Tehran with professionally confirmed infestation. The findings showed that participants had modest knowledge of German cockroach biology and infestation pathways, alongside intermediate levels of preventive behavior. Among the examined Health Belief Model constructs, knowledge, perceived benefits, and self-efficacy were independently associated with preventive practices and demonstrated modest predictive value within the regression model. These results suggest that risk awareness alone may be insufficient to promote sustainable household pest-control practices. Instead, interventions should prioritize improving factual knowledge, strengthening beliefs in the effectiveness of preventive measures, and enhancing residents’ confidence in implementing control behaviors.

Given the modest explanatory power of the regression model, preventive practices are likely influenced by broader environmental, structural, and socioeconomic factors beyond individual cognition. Future urban cockroach-management programs should therefore integrate behavioral education with housing improvement, environmental sanitation, and accessible pest-control services to achieve more sustainable infestation control.

## Data Availability

All data produced in the present study are available upon reasonable request to the authors

## Declarations

### Ethics Approval and Consent to Participate

The study protocol was approved by the Ethics Committee of Shiraz University of Medical Sciences (Approval Code: IR.SUMS.SCHEANUT.REC.1405.013) Written informed consent was obtained from all participants.

### Consent for Publication

Not applicable.

### Availability of Data and Materials

The datasets generated and analyzed during the current study are available from the corresponding author upon reasonable request.

## Authors’ Contributions

Conceptualization: S.M., A.A., M.D.M-F.; Methodology: S.M., A.A., H.G.M.; Data Collection: S.M., S.B.; Formal Analysis: S.M., M.K., H.G.M.; Writing—Original Draft: S.M.; Writing—Review & Editing: All authors; Supervision: M.D.M-F. All authors approved the final manuscript.

## Acknowledgments

The authors would like to thank all participants and pest-management professionals who assisted with household inspections and data collection. Our sincere gratitude goes to the Vice-Chancellorship for Research and Technology at Shiraz University of Medical Sciences for the approval of this project. We are also indebted to our colleagues at Research Center for Health and Sciences for processing this proposal.

