## Supplemental Table for "Behavioral determinants of preventive practices against German cockroach infestation among urban residents in Tehran, Iran"

**Supplementary Materials**

**Table S1. Item-level responses for perceived susceptibility related to household German cockroach infestation among urban residents in Tehran, Iran.**

| Code | Item | Mean ± SD | Min | Max |
| --- | --- | --- | --- | --- |
| Q17 | Based on the sanitation condition of our home, there is a possibility of cockroach infestation | 2.792 ± 1.321 | 1 | 5 |
| Q18 | Due to the age of our building, there is a possibility of cockroach infestation | 3.275 ± 1.223 | 1 | 5 |
| Q19 | Due to the environmental conditions of my neighborhood, cockroach infestation is likely to occur | 3.067 ± 1.200 | 1 | 5 |

**Footnote:** Response scale: 1 = Strongly disagree, 5 = Strongly agree.

**Table S2. Item-level responses for perceived severity related to household German cockroach infestation among urban residents in Tehran, Iran.**

| Code | Item | Mean ± SD | Min | Max |
| --- | --- | --- | --- | --- |
| Q20 | Cockroaches can transmit diseases to humans | 3.508 ± 1.077 | 1 | 5 |
| Q21 | Presence of cockroaches at home may cause embarrassment in front of guests | 3.125 ± 1.254 | 1 | 5 |
| Q22 | Cockroaches can contaminate food materials | 3.683 ± 1.130 | 1 | 5 |
| Q23 | Cockroach infestation at home is common and not important | 2.600 ± 1.155 | 1 | 5 |

**Footnote:** Response scale: 1 = Strongly disagree, 5 = Strongly agree. Higher scores indicate greater perceived severity.

**Table S3. Item-level responses for perceived benefits of German cockroach control measures among urban residents in Tehran, Iran.**

| Code | Item | Mean ± SD | Min | Max |
| --- | --- | --- | --- | --- |
| Q24 | Household cleaning and washing clothes are useful for cockroach control | 3.725 ± 1.084 | 1 | 5 |
| Q25 | Discarding old household items is useful for cockroach control | 3.100 ± 1.191 | 1 | 5 |
| Q26 | Use of poisoned bait is useful for cockroach control | 3.367 ± 1.276 | 1 | 5 |
| Q27 | Household insecticide spraying is useful for cockroach control | 3.525 ± 1.195 | 1 | 5 |

**Footnote:** Response scale: 1 = Very low, 5 = Very high.

**Table S4. Item-level responses for perceived barriers to German cockroach control among urban residents in Tehran, Iran.**

| Code | Item | Mean ± SD | Min | Max |
| --- | --- | --- | --- | --- |
| Q28 | Cockroach control measures are time-consuming and exhausting | 3.033 ± 1.236 | 1 | 5 |
| Q29 | Cockroach-control products and equipment are expensive | 2.983 ± 1.202 | 1 | 5 |
| Q30 | Professional pest-control consultation costs are high | 2.775 ± 1.240 | 1 | 5 |

**Footnote:** Response scale: 1 = Strongly disagree, 5 = Strongly agree. Higher scores indicate greater perceived barriers.

**Table S5. Item-level responses for self-efficacy related to German cockroach control among urban residents in Tehran, Iran.**

| Code | Item | Mean ± SD | Min | Max |
| --- | --- | --- | --- | --- |
| Q31 | Despite the difficulty of cockroach control, I can perform preventive measures | 3.250 ± 1.238 | 1 | 5 |
| Q32 | I can afford cockroach-control products and equipment | 2.983 ± 1.322 | 1 | 5 |
| Q33 | I can afford professional pest-control consultation costs | 2.783 ± 1.271 | 1 | 5 |
| Q34 | I can convince myself to discard old and unnecessary household items | 3.308 ± 1.275 | 1 | 5 |

**Footnote:** Response scale: 1 = Very low, 5 = Very high. Higher scores indicate greater self-efficacy.

**Table S6. Item-level responses for preventive and control practices related to household German cockroach infestation among urban residents in Tehran, Iran.**

| Code | Item | Mean ± SD | Min | Max |
| --- | --- | --- | --- | --- |
| Q35 | Household cleaning and washing clothes | 3.475 ± 1.243 | 1 | 5 |
| Q36 | Discarding or destroying old household items | 2.808 ± 1.349 | 1 | 5 |
| Q37 | Use of poisoned bait | 3.150 ± 1.281 | 1 | 5 |
| Q38 | Household insecticide spraying | 3.375 ± 1.174 | 1 | 5 |
| Q39 | Consulting professional pest-control specialists | 2.975 ± 1.233 | 1 | 5 |

**Footnote:** Response scale: 1 = Never/Very low, 5 = Always/Very high. Higher scores indicate more frequent engagement in preventive practices.
